# Low-Cost Precision nutrition recommendations, generated by metataxonomy-based microbiome tests, improve food group choices and gut health indicators in a population with obesity diagnosis in Colombia

**DOI:** 10.64898/2026.04.28.25331845

**Authors:** Vanesa Caro Miranda, Shadia Blel Jubiz, Isabel Adarve-Rengifo, Sara Londoño-Osorio, Maria Clara Arrieta-Echeverri, Laura Gómez-Mesa, Juan David Serna Tangarife, Álvaro Muñoz, Carlos Andrés Zapata, Laura Sierra-Zapata

**Affiliations:** Universidad EAFIT, Natural Systems and Sustainability Area; School of Applied Sciences and Engineering, Medellín, Colombia; Astrolab Biotecnología SAS, Medellín, Colombia; COMFAMA, Family compensation fund from Antioquia, Medellín, Colombia; Cazamed, Medicina interna y hábitos saludables

**Keywords:** : Gastrointestinal microbiome, Obesity, Precision Nutrition, Dietary Intake, Cardiometabolic health, Colombia and Latin America

## Abstract

**Aims:** This study aimed to explore the relationship between gut microbiota composition, obesity, and the effects of a dietary intervention in 50 participants with obesity diagnosis from Antioquia, Colombia.

**Methods:** A single-blind intervention study was conducted, with 25 participants assigned to a control group (CG) and 25 to an intervention group (IG), these last followed a microbiota-enhancing dietary plan for 90 consecutive days. Gut microbiota changes were assessed by sequencing region V3-V4 of 16S rRNA gene and applying the analytical methodology of Biomatest® gut health index. Blood biomarkers, including HbA1C, cholesterol, HDL, LDL, triglycerides, and glucose, were measured at baseline and post-intervention.

**Results:** *Prevotella* and *Succinivibrio* were prevalent in the study population. The IG showed significant increases in gut microbial diversity (Shannon index) from baseline to post-intervention. Both groups exhibited significant changes in the Biomatest gut health index, with significant improvements in the IG. Significant correlations were found between dietary intake, blood biomarkers, and microbial abundances, such as the direct association between serum glucose and ultra-processed food intake and between total cholesterol and *Dialister*. Fish and seafood consumption correlated positively with *Akkermansia*, while egg intake was associated with higher levels of *Desulfovibrio,* and *Lactobacillus* with decreased glycated hemoglobin. The IG experienced a significant rise in *Roseburia*, a gut health biomarker, while the CG showed higher levels in inflammatory groups like *Fusobacteriota*.

**Conclusions:** Dietary intake significantly influences gut microbiota composition and blood biomarkers. Nutritional programs that improve gut microbiota, as demonstrated by the IG, positively impact gut health in people with obesity diagnosis and may influence healthier dietary choices. These findings support integrating microbiota diagnostics into personalized nutrition strategies, contributing valuable data on Latin American populations.

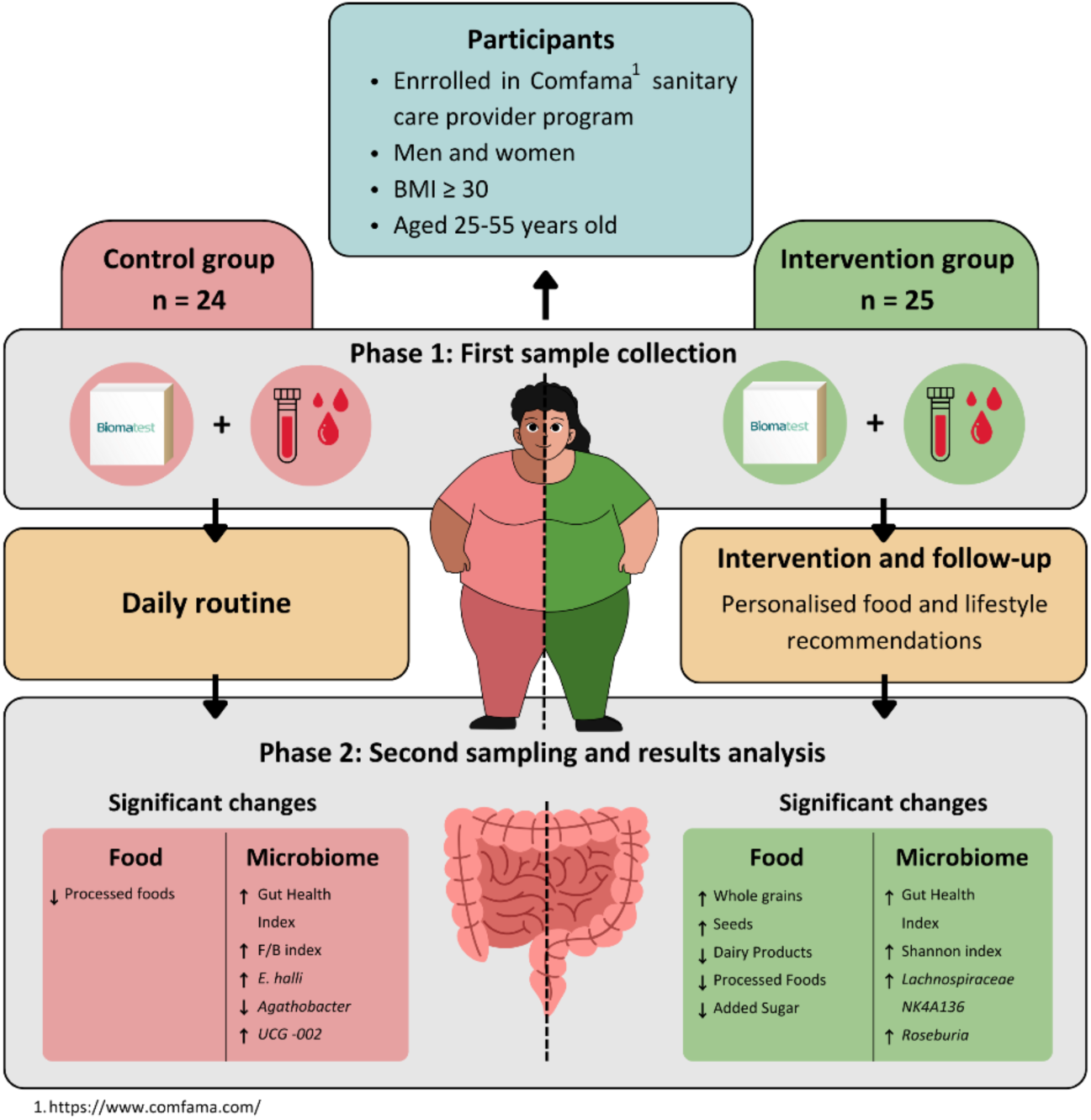

## Introduction

The ecosystem of microorganisms residing in the gastrointestinal tract, known as the gut microbiota or microbiome—the latter term being more inclusive as it encompasses the metabolites and genetic material of this community^1^ has been shown to be a fundamental component in regulating various physiological processes, such as the breakdown of indigestible polysaccharides,^2^ the formation of the intestinal epithelium, the management of dietary energy intake, and nutrient absorption in the intestine.^3, 4, 5, 6^ In the field of metabolic health, the gut microbiome is a key point of scientific research, with its composition and functionality intrinsically linked to the development and progression of conditions like obesity.^7, 8, 9^ Recent evidence has also highlighted the potential benefits of regulating or restoring this ecosystem to a healthy state (a term known as eubiosis),^10, 11^ leading to a reduction in chronic inflammation and metabolic syndrome.^12, 13^ Beyond its impact on physical health, there have been findings linking the composition of the gut microbiota to neuropsychiatric disorders, including depression and anxiety.^14, 15, 16, 17^ Although the precise mechanisms of this interaction are still being uncovered, there is an urgent need to explore the holistic impact of the microbiome on human health. The understanding of its role has evolved from a primary focus on digestion to broader considerations of its influence on immune response and metabolism.

Pioneering studies have revealed distinctive microbial signatures associated with obesity in Western populations, linked to a decrease in bacteria considered beneficial for health, such as the genera *Bacillus* sp., *Lactobacillus* sp., *Enterococcus*, *Clostridium*, *Ruminococcus*, *Roseburia*, *Faecalibacterium*, *Coprococcus* sp., *Eubacterium* sp., *Oscillibacter* sp., *Prevotella* sp.^18, 19, 20, 21, 22^ This has prompted a paradigm shift in our understanding of the relationship between microbial communities and metabolic status. It has also been widely described that obesity is associated with gut dysbiosis (low diversity indices in the gut, prevalence of inflammatory and aerobic bacteria) and a high Firmicutes/Bacteroidetes ratio.^23^

A recent study presented by Spanish researcher Paula Aranaz from the Navarra Institute for Health Research at the 2024 European Congress on Obesity showed that certain specific bacteria are predictors of obesity, but they are involved differently in men and women.^24^ The study found that people in the high-obesity index group had low levels of *Christensenella minuta*, a bacterial species associated with leanness and good health. Significant differences were also found in the microbial species present in the gut of men and women with obesity. Men in the high-obesity index group had higher levels of *Parabacteroides helcogenes* and *Campylobacter canadensis*, while women in this group had higher levels of three *Prevotella* species: *Prevotella micans*, *Prevotella brevis*, and *Prevotella sacharolytica*. These species were associated not only with higher BMI but also with greater body fat and abdominal circumference. By performing untargeted metabolic analysis on plasma samples from the study participants, the researchers found that the levels of phospholipids and sphingolipids differed between participants with high and low obesity indices. Both bioactive lipids are associated with metabolic diseases such as diabetes and related vascular complications. Another study found that the novel strain DSM33407 of *Christensenella minuta* may act as an anti-obesogenic agent in preclinical murine models.^23^

Various studies continue to demonstrate sex-specific patterns in the gut microbiome of people with obesity diagnosis, with these differences even being evident during the reproductive stage of women. In the study conducted by Haro et al. (2016),^25^ it was observed that the abundance of the genus *Bacteroides* was lower in men with obesity diagnosis than in women with obesity diagnosis. The study also found a higher presence of the genera *Veillonella* and *Methanobrevibacter* in men compared to women.

Although these studies have improved our understanding of the relationship between the gut microbiome and obesity, it is crucial to recognize the need for region-specific research, as microbial compositions can vary considerably based on geographic location, lifestyle, and ethnicity.^26, 27, 28^ This reality motivated our investigation in the Colombian population, a demographic group where the interaction between the gut microbiome and obesity remains a relatively unexplored field. Research on obesity in Colombia has shown that a lower BMI is associated with the presence of primary fiber degraders, and these bacteria impact the host’s energy balance.^8^ So far, it has been found that the gut microbiome of Colombians differs from that of Americans, Europeans, and Asians in terms of variations related to the low intake of fats and total proteins and the high intake of carbohydrates and fibers. Additionally, a shift in the Firmicutes and Bacteroidota was found in cases of obesity, as there is a tendency in Colombians for Firmicutes to decrease with increasing BMI, while no changes were observed in Bacteroidota.^29^

Colombia, despite its growing prevalence of obesity (36% of the population is overweight and 21% has obesity),^30, 31^ presents a unique context influenced by a confluence of genetic, dietary, and environmental factors. However, the research landscape in Colombia regarding the interaction between the gut microbiome and obesity is notably scarce. The lack of specific studies addressing this relationship within the Colombian context underscores the importance of this research in helping to close this knowledge gap. This study, which includes a cohort of 50 participants with obesity diagnosis enrolled with the healthcare provider COMFAMA in Antioquia, Colombia, aims to describe the relationship between the gut microbiome and obesity within this Latin American population. This approach aligns with the broader scientific goal of examining microbial signatures associated with obesity across diverse ethnic groups, moving beyond the predominantly Western focus.

Traditionally, obesity is addressed through lifestyle changes, nutritional education, and modification and increase of physical exercise, all of which are fundamental for the long-term success of treatment. Other treatments include anorexigenic medications, very low-calorie diets, and surgical techniques, which may be necessary and have a clinical role in certain groups of patients with extreme obesity and cardiovascular complications, according to the classification levels of obesity: class 1: 30–35 kg/m² (BMI); class 2: 35–40 kg/m²; class 3: >40 kg/m². However, the prevention of obesity is considered a highly important strategy for societies, and large-scale population studies have shown that it is possible to modify behavior and reduce cardiovascular risk.^32, 33^ These habit-change strategies in dietary terms generally include calorie restriction, the reduction of ultra-processed and calorie-dense foods; the reduction of fat intake, and, in contrast, an increase in fruits and vegetables, plant-based diets, whole and unrefined grains, and plant proteins.^34^ These dietary plans did not emphasize increasing fiber and restoring the balance of the gut microbiome (eubiosis), and it is only in recent years that this has begun to be considered an important marker for adjusting these treatments. It is expected that research in this area will continue to contribute, soon, more creative and inclusive therapies and tools emerging from multidisciplinary teams of doctors, nutritionists, exercise physiologists, psychologists, and other disciplines.^32^

This research adopts a holistic perspective, aiming to discern whether the microbial signatures associated with obesity in Western contexts hold true for the Colombian population, whether distinct microbial profiles contribute to the obesity landscape in this region, and whether, through microbiome diagnosis and dietary plan adjustments aimed at restoring it, favorable changes can be generated in patients with this condition. The core of this study involves a central hypothesis focused on observing a positive change in participants who follow the prescribed diet or lifestyle compared to those who maintain their regular routines. This goal highlights our commitment not only to unravel the relationship between the gut microbiome and obesity but also to assess the real-world impact of dietary and lifestyle interventions on microbial dynamics and, consequently, metabolic health. By delivering these results, we not only expand the knowledge of the global variability of the microbiome but also pave the way for implementing more precise strategies in the prevention and treatment of obesity in specific contexts.

## Methodology

### Ethical Considerations

This study was approved by the ethics committee of Universidad EAFIT, established by Act No. 03-89-1109-2012 of the Research Committee on September 11, 2012, and created as the Institutional Ethics Committee through Acts 457 of October 1, 2014, and 474 of April 26, 2017, by the University’s Governing Board (Supplementary Material 1). Each volunteer signed an informed consent form to authorize the processing of the samples and data before enrolling in the study. All participants were informed about the procedures of the study. Additionally, participants were assured anonymity and confidentiality of the collected data.

### Study Design

This exploratory study was designed as a simple, blinded interventional study. The cohort included 50 participants enrolled in the COMFAMA healthcare provider, diagnosed with obesity, defined as a BMI ≥30. They were divided into two groups: 25 in the IG and 25 in the control CG.

### Selection and Recruitment of Participants

The study consisted of three distinct phases: (i) participant recruitment phase; (ii) Phase 1, consisted of the initial collection of blood and gut microbiome samples, analysis, and delivery of results to the participants; the intervention and follow-up period (90 days), and (iii) Phase 2, which corresponded to the second collection of blood and gut microbiome samples, analysis, and delivery of results to the participants.

Participant Recruitment Phase. For the participants selection, a database from the healthcare provider COMFAMA was used, containing both men and women who initially met the diagnostic criterion for obesity, as defined by the World Health Organization (WHO), of a BMI greater than or equal to 30, and considering the inclusion and exclusion criteria (Supplementary Material 2). These identified users were contacted by phone to provide detailed information about the study, the conditions for participation, and the sample collection procedure. People who declined to participate or did not meet the criteria were excluded, and recruitment continued until a confirmed base of 50 participants was reached.

Phase 1: Initial Sample Collection. Confirmation of participation during this phase was crucial to ensure the full commitment of participants and their understanding of the study’s objectives and procedures.

For the gut microbiome analysis, Biomatest, led by Astrolab Biotecnología (hereinafter Astrolab Bio), sent each participant a sample collection kit to their home along with the informed consent form (Supplementary Material 3) for them to sign. The samples were collected and safely transported to the laboratory for processing.

To obtain a more comprehensive metabolic health profile of the participants, anthropometric data (weight, height) and blood samples were collected to measure a full lipid profile (total cholesterol, triglycerides, HDL, and LDL) and blood glucose levels (HbA1c). This process was handled by COMFAMA personnel assigned to the project.

The delivery of the gut microbiome diagnostic test results from this phase took place at COMFAMA’s facilities with each group separately, maintaining the single-blind strategy for the participants. Likewise, the 50 participants had access to medical support for reviewing the results of the blood tests, and only the 25 in the IG received support from COMFAMA nutrition specialists to emphasize and monitor the recommendations to improve gut microbiome health provided by Biomatest.

### Report of Gut Microbiome Results from Biomatest Based on Metataxonomy

The gut microbiome results generated for everyone in the study were translated into actionable indicators, based on the relative abundance of each of the bacterial taxa present in the gut and their effects on human health. These are determined according to a proprietary and secret methodology developed by Astrolab Bio (protected by a trade secret owned by Universidad EAFIT, Supplementary Material 4). It mainly consists of describing the composition of the gut bacterial community and reporting indices such as the Firmicutes/Bacteroidota ratio (F/B ratio), top 15 “beneficial” and “pro-inflammatory” bacteria, gut enterotype, ecological diversity indices, and the Biomatest gut health index. All of this is translated into personalized effects and interpretations based on everyone’s metadata, ultimately providing actionable recommendations focused on diet and lifestyle.

### Design of the Precision Nutrition Intervention Based on the Gut Microbiome

The development and design of the dietary intervention involved the participation of the medical team allied with Astrolab Bio, consisting of a nutritionist-dietitian and an internist, and the Astrolab Bio team, ensuring an integrative and scientifically sound approach. This intervention primarily focused on the inclusion of food groups rather than macronutrients, with the goal of simplifying dietary recommendations and improving participant adherence. These food groups were selected for their beneficial impact on gut microbiome health in cases where dysbiosis in the gut microbiome is expected (as we hypothesize may be the case for the study population): legumes, whole grains, tubers, fruits, vegetables, nuts, and seeds. These foods are rich in fiber and phytonutrients, which are essential for the proliferation and growth of beneficial bacteria in the gut. Furthermore, it is important to note that this intervention did not focus on restricting the consumption of ultra-processed, processed foods, saturated fats, canned goods, refined sugar, and others, but instead recommended significantly reducing such foods and including and diversifying the recommended foods.

Participants were also provided with a “rainbow chart” to exemplify the various colors of the foods they should include in their diet. This approach was based on the principle that a colorful plate often indicates a nutritionally balanced and healthy diet, rich in different nutrients and antioxidants that improve gut microbiome diversity and balance. By encouraging the consumption of a variety of colorful foods, the goal was to enhance the overall appeal and health benefits of dietary intervention.

Because diet constitutes the main modulating factor of the gut microbiota in both the short and long term, we proposed a dietary intervention adapted to the cultural and socioeconomic context of a low-income country. The aim is to ensure the long-term sustainability of the dietary pattern and to evaluate its impact on the gut microbiota.

Our dietary intervention is based on an accessible food model aligned with local customs and preferences, prioritizing the consumption of whole foods from agriculture. We have designed a dietary scheme centered on plant sources, including legumes, whole grains, fruits, seeds and specific tubers such as potatoes, which have been shown to exert a beneficial influence on the intestinal microbiota.^35^ At the same time, the consumption of foods characteristic of the Western or urban dietary pattern, which is characterized by a high intake of processed products, added sugars and foods rich in saturated animal fats, as well as limited dietary diversity, has been reduced. ^36, 37^

In summary, our intervention promotes a dietary pattern closer to that of rural communities than that of urban populations, which has been evaluated in several studies with evident benefits on the intestinal microbiota.^38^ This food model, based on local agriculture, is not only accessible to low-income populations, but also favors daily dietary diversity. As a result, it complies with the fundamental principles of an optimal diet for the intestinal microbiota: rich in whole foods, predominantly of plant origin, varied and high in fiber, with a reduced intake of aggressive factors such as saturated fats from animal foods and ultra-processed products, whose thermal processing can negatively impact the intestinal environment.^35^

Dietary patterns such as the one implemented in our study induce rapid changes in the composition of the gut microbiota in the short term and, in the long term, generate favorable modifications in the genomic structure and metabolic functionality of the microbiota.^38, 39^

### Intervention and Follow-up Phase

After the sample collections and the initial delivery of Biomatest gut microbiome analysis, a 90-day period began during which participants in the IG followed the nutrition and lifestyle recommendations designed by the Astrolab Bio team, focused on gut microbiome modulation. Meanwhile, the participants in the CG received both gut microbiome and blood results but did not receive specific nutrition and lifestyle recommendations, allowing them to maintain their usual routine. This strategy allowed for a meaningful assessment of the impact of the proposed interventions. To monitor participants during this intervention period, a detailed questionnaire (Supplementary Material 5) was designed and provided to both the intervention and CG, allowing for the visualization of study adherence, key dietary habits, and some aspects of the participants’ mental health. This questionnaire was sent every 15 days via WhatsApp, totaling 6 questionnaires per person. Additionally, a WhatsApp group was created for the IG, and weekly graphic materials were shared, related to the benefits and recipes of the recommended food groups.

Phase 2: Second Sample Collection and Analysis of Project Results. At the end of the intervention phase, the Biomatest gut microbiome samples were collected again, along with anthropometric data (weight, height) and blood samples. Likewise, the 50 participants received medical and nutritional support from COMFAMA based on the final nutrition and lifestyle recommendations determined by the project. The delivery of gut microbiome results took place in person at COMFAMA’s facilities for each study group. In this phase, both gut microbiome and blood results were analyzed and compared with those taken in Phase 1 to measure the effects of the personalized recommendations on the gut microbiome status of this group of participants.

### Collection and Processing of Gut Microbiome Samples

Gut microbiome data collection was performed using the Biomatest at-home sample collection kit provided by Astrolab Bio.^40^ This kit includes a collection tube containing an aqueous solution that ensures the stability of fecal samples at room temperature, allowing participants to easily collect the sample in the comfort of their homes. In addition, the kit provides complete instructions to guide users through the sample collection process.

The DNA extraction from the fecal samples was performed using Norgen’s Stool DNA Isolation kit (Cat. 27600), following the manufacturer’s protocol and ensuring high-quality genetic material for subsequent analysis. The V3-V4 region of the 16S rRNA gene was sequenced on the Illumina MiSeq and Novaseq6000 platforms. All collected samples underwent taxonomic classification, and their Amplicon Sequence Variants (ASV) were analyzed. The raw data files (.fastq) were processed in QIIME2, using deblur for demultiplexing, consensual Blast, and the SILVA database (SSU 138.1 version) for taxonomic classification.^41^ The bioinformatics workflow allowed for the calculation of the abundance of each taxon in the participants’ gut microbiomes.

### Data and Statistical Analysis

All statistical analyses were performed using the R statistical software version 4.3.2.^42^ANOVA test was conducted for normally distributed data, and the non-parametric Kruskall-Wallis test was used for those not meeting normality, to compare the study groups and within each group, considering body composition (weight, BMI, height), metabolic parameters (blood chemistry), and the various metrics that make up the Biomatest gut microbiome report (alpha diversity, gut health index, F/B ratio, and relative abundance of bacterial groups). Average differences were considered statistically significant when p ≤ 0.05.

The figures presented in this study were generated using Microsoft PowerBI data visualization software. The percentage change (Δ) in food consumption was calculated using the following equation:

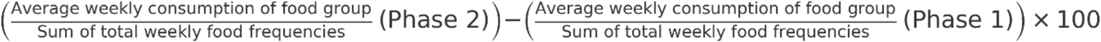

For some of the visualizations, error bars were incorporated to represent data variability and provide a better understanding of confidence intervals. These error bars were calculated using the standard deviation (SD) divided by the square root of the number of data points (n), thus providing a visual representation of the standard error of the mean.

For correlation analyses between the relative abundance of gut bacteria, dietary data, and metabolic parameters, the R package corrplot was used.^43^ Correlations were calculated using Pearson’s correlation coefficient, and the resulting graphs provided a clear and understandable representation of the interactions between the various variables studied. Specifically, correlations were analyzed based on the delta of each measured variable, meaning the change between phase 2 and phase 1. This approach allowed us to assess whether the variables increased or decreased in relation to other correlated variables. Only correlation values greater than or equal to 0.4 and less than or equal to -0.4 were considered significant, to highlight the most relevant relationships.

## Results

### Recruitment Phase

Of the 50 participants contacted to evaluate the effect of nutritional recommendations on improving the gut microbiome in a cohort of obesity diagnosis, 24 participated in the groupCG and 25 in the IG. The difference in group sizes was due to one participant withdrawing during Phase 1 of the study.

The main characteristics of the participants are shown in Table 1, highlighting that the majority were women (83.4% in the CG and 80% in the IG), with an average BMI of 36 kg/m² in both groups, and an average age of 36 years. Information on blood chemistry is also provided for both study phases in the CG and IG.

**Table 1.** Characteristics of participants in Phase 1 and Phase 2 of the study. . The sample comprises n=25 participants in the IG and n=24 in the CG. Results are expressed as the mean along with the corresponding standard deviation (X ± SD). Statistical significance is indicated by the *p-value*, calculated using one-way ANOVA for pairwise comparisons.

| Characteristics/Parameters | Intervention group<br>$\bar{X} \pm SD$ | Control group<br>$\bar{X} \pm SD$ | *p-value* |
| --- | --- | --- | --- |
| **Sex (%)** |  |  |  |
| Female | 80 | 83,4 |  |
| Male | 20 | 16,6 |  |
| **Age (years)** | 36,04 ± 7,04 | 36,91 ± 7,40 | 0.68 |
| **Height (cm)** | 162,3 ± 0,08 | 162,6 ± 0,06 | 0.89 |
| **Body mass (kg)** |  |  |  |
| Phase 1 | 95,50 ± 17,17 | 97,44 ± 14,63 | 0.68 |
| Phase 2 | 93,65 ± 19,22 | 96,26 ± 14,53 | 0,60 |
| Body mass change (%) | -6,63 ± 20,82 | -1,06 ± 6,26 | 0,27 |
| **BMI (kg/m<sup>2</sup>)** |  |  |  |
| Phase 1 | 36,11 ± 4,74 | 36,87 ± 4,60 | 0.58 |
| Phase 2 | 35,34 ± 5,34 | 35,08 ± 9,12 | 0,44 |
| BMI change (%) | -10,98 ± 28,42 | -5,67 ± 21,73 | 0,50 |
| **Serum Glucose (mg/dL)** |  |  |  |
| Phase 1 | 94,11 ± 11,46 | 89,13 ± 8,52 | 0,17 |
| Phase 2 | 96,47 ± 16,09 | 88,4 ± 8,20 | 0,05 |
| Serum Glucose change (%) | 2,57 ± 10,06 | -0,13 ± 11,59 | 0,48 |
| **HbA1C (%)** |  |  |  |
| Phase 1 | 0,05 ± 0,003 | 0,05 ± 0,002 | 0,19 |
| Phase 2 | 0,05 ± 0,003 | 0,05 ± 0,003 | 0,18 |
| HbA1C change (%) | 0,32 ± 3,20 | -0,21 ± 4,78 | 0,74 |
| **Total cholesterol (mg/dL)** |  |  |  |
| Phase 1 | 189,70 ± 21,95 | 182,2 ± 28,61 | 0,40 |
| Phase 2 | 189,88 ± 14,72 | 180,33 ± 36,63 | 0,35 |
| Total cholesterol change (%) | 0,80 ± 8,69 | -0,31 ± 15,87 | 0,83 |
| **HDL (mg/dL)** |  |  |  |
| Phase 1 | 44,41 ± 9,32 | 43,33 ± 9,30 | 0,74 |
| Phase 2 | 45,58 ± 8,28 | 44,06 ± 10,66 | 0,63 |
| HDL change (%) | 3,91 ± 11,98 | 1,32 ± 8,36 | 0,65 |
| **LDL (mg/dL)** |  |  |  |
| Phase 1 | 117 ± 24,02 | 109,58 ± 22,73 | 0,37 |
| Phase 2 | 119,21 ± 17,87 | 111,38 ± 31,46 | 0,47 |
| LDL change (%) | 4,02 ± 13,39 | 1,69 ± 18,23 | 0,68 |
| **Triglycerides (mg/dL)** |  |  |  |
| Phase 1 | 141,47 ± 87,41 | 146,4 ± 66,50 | 0,70 |
| Phase 2 | 125,41 ± 82,04 | 124,4 ± 47,89 | 0,36 |
| Triglycerides change (%) | -5,97 ± 24,91 | -5,26 ± 28,84 | 0,94 |

**Abbreviations:** BMI, body mass index; HDL, high-density lipoprotein; LDL, low-density lipoprotein; **HbA1C**, glycated haemoglobin.

### Study Adherence

Overall, 92% of adherence to the study was observed in terms of the gut microbiome testing, with 23 participants in each group (control and intervention) participating in phase 2. Similar results were noted regarding metadata questionnaire responses, which were crucial for analyzing and linking the gut microbiota results.However, blood chemistry data collected through blood tests at the sampling points of Sura - COMFAMA posed a challenge; there was lower adherence in this parameter, with 18 participants in the CG (72%) and 17 in the IG (68%) completing the blood tests in phase 2. For the Metadata Questionnaire, the adherence percentage in the CG was **95.83%** in Phase 2, compared to 100% in Phase 1. In the IG, adherence was **92%** in Phase 2, down from 100% in Phase 1.

### Dietary Analysis

The dietary analysis sets the stage for correlating changes in the microbiome with specific dietary habits, providing a more holistic perspective on the interaction between diet and gut microbiome composition in the context of obesity. Dietary patterns of the participants were assessed through a comprehensive survey; in the initial questionnaire, participants were asked about their weekly consumption of specific food categories, which provided insights into their dietary preferences. This information is presented in Figure 1.

**Figure 1.**
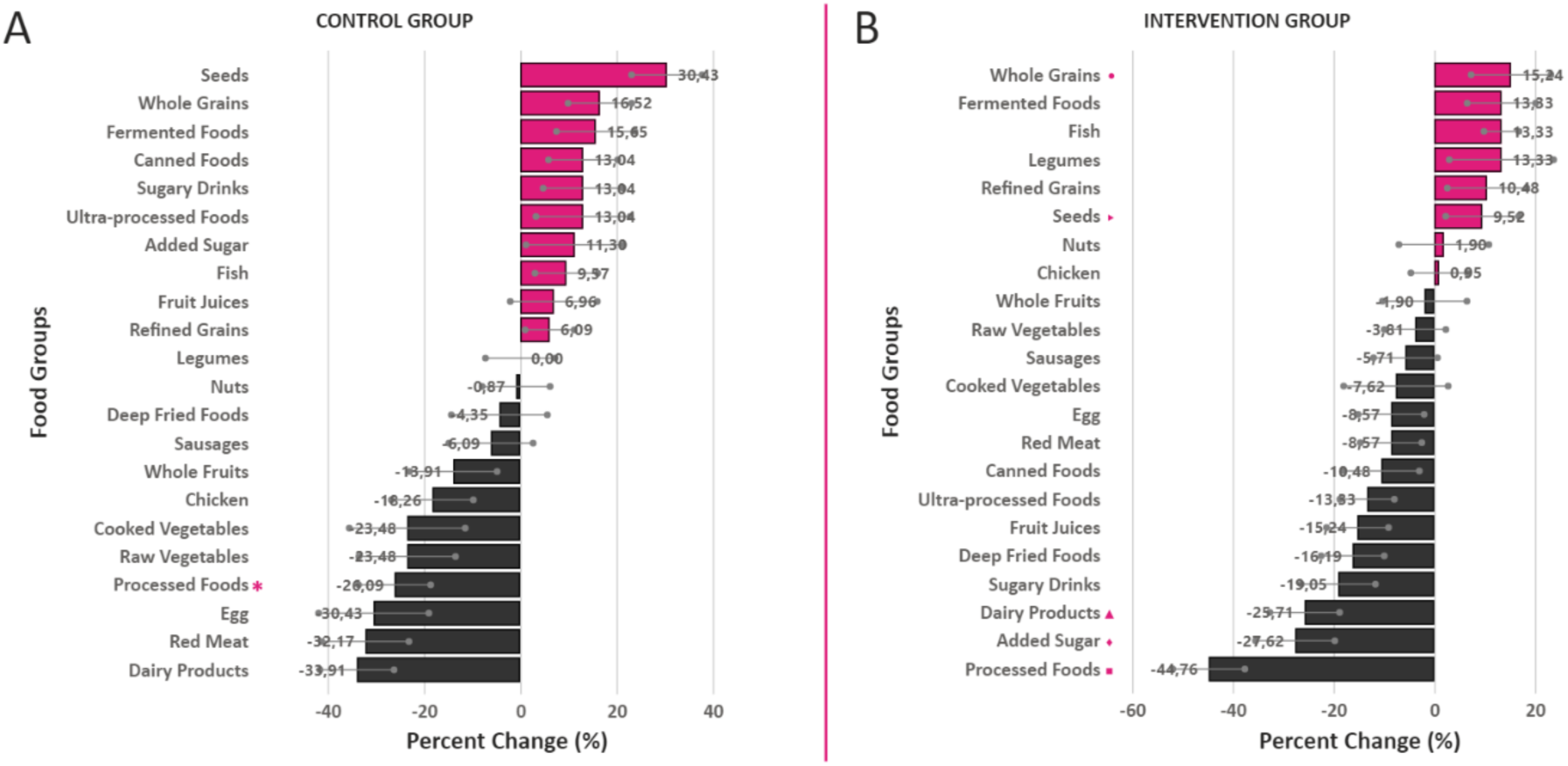
Changes in food group consumption throughout the study. (A) Changes within the CG and (B) changes within the IG (n = 46). Food groups with statistically significant changes are marked with pink asterisks, as determined by a nonparametric Kruskal-Wallis analysis at a significance level of **p < 0.05**.

### Body Composition and Metabolic Parameters

No statistically significant differences were observed in body composition parameters (body mass, BMI) or obesity-related blood biomarkers (serum glucose, HbA1C, total cholesterol, HDL, LDL, triglycerides) between the CG and IG in both phases of the study (Table 1).

### Lifestyle comparison of participants in phase 1 and phase 2 of the study

The demographic analysis of habits and behavioral data throughout the study are detailed in Figure 1. Regarding the consumption of tobacco, most participants in both groups and phases reported abstinence (Fig. 1A). Regarding alcohol consumption, the CG in phase 1 presented the highest consumption, however, consumption decreased from 13 to 10 participants, while in the intervention group a decrease in alcohol consumption was observed from 8 to 4 (Fig. 1B). Regarding physical exercise, there was an increase in the number of participants who did not exercise in both groups: in the intervention group from 12 to 13 and in the CG from 16 to 18 (Fig. 1C). Regarding sleep, most participants in both groups and phases reported sleeping an average of 6 to 8 hours per day (Fig. 1D).

Regarding the observed health conditions (Fig. 1E), they were identified and classified into ten distinct categories: metabolic diseases, gastrointestinal diseases, thyroid gland diseases, fatty liver, anemia, polycystic ovary, dermatological diseases, mental diseases, respiratory diseases, and fibromyalgia. Within the IG, the most prevalent conditions were gastrointestinal diseases (n=6), followed by dermatological diseases (n=3) and mental diseases (n=2). On the other hand, in the CG, the most prevalent conditions were gastrointestinal diseases (n=3) and metabolic diseases (n=3).

In the control group, significant differences were observed in relation to processed food intake (*p-value = 0.04) when comparing Phase 1 with Phase 2. In the IG, significant differences were observed in the consumption of dairy (▴p-value = 0.01), processed foods (▪p-value = 0.00), added sugar (◆p-value = 0.03), whole grains (● p-value = 0.00) and seeds (►p-value = 0.01) when comparing Phase 1 with Phase 2. The analysis takes a deep approach into the frequency of key dietary components, allowing for precise understanding of the participants’ nutritional intake.

### Impact of the Intervention on Gut microbiome Indicators

The impact of personalised recommendations on the gut microbiome composition of study participants, before and after the intervention, is evidenced in Figure 2. In these figures, significant differences can be observed for various indicators. In the CG (Fig. 2A and 2B), where no personalised recommendations based on microbiome composition were given, an increase in the phyla Actinobacteriota, Desulfobacteriota, Verrucomicrobiota, Fusobacteriota, and the genera UCG-002 and *Eubacterium halii* was observed. Likewise, in this group, there was a decrease in the phylum Bacteroidota and the genus *Agathobacter*. In the IG, where personalised recommendations based on gut microbiome composition and follow-up were provided, a significant increase in the phylum Firmicutes and the genera Lachnospiraceae NK4A 136 group and *Roseburia* was observed (Fig. 2C and 2D). The gut health index used by the employed test (see calculation in methods) increased significantly in both study groups (Fig. 2E); however, diversity increased exclusively in the IG (Fig. 2F), and the F/B ratio increased significantly in the CG (Fig. 2G). *Prevotella* was the most abundant genus in both phase 1 and phase 2 of the intervention and CG. Furthermore, correlations were found between gut microbiome biomarkers, blood chemistry biomarkers, and changes in food consumption habits (Figure 3).

**Figure 2.**
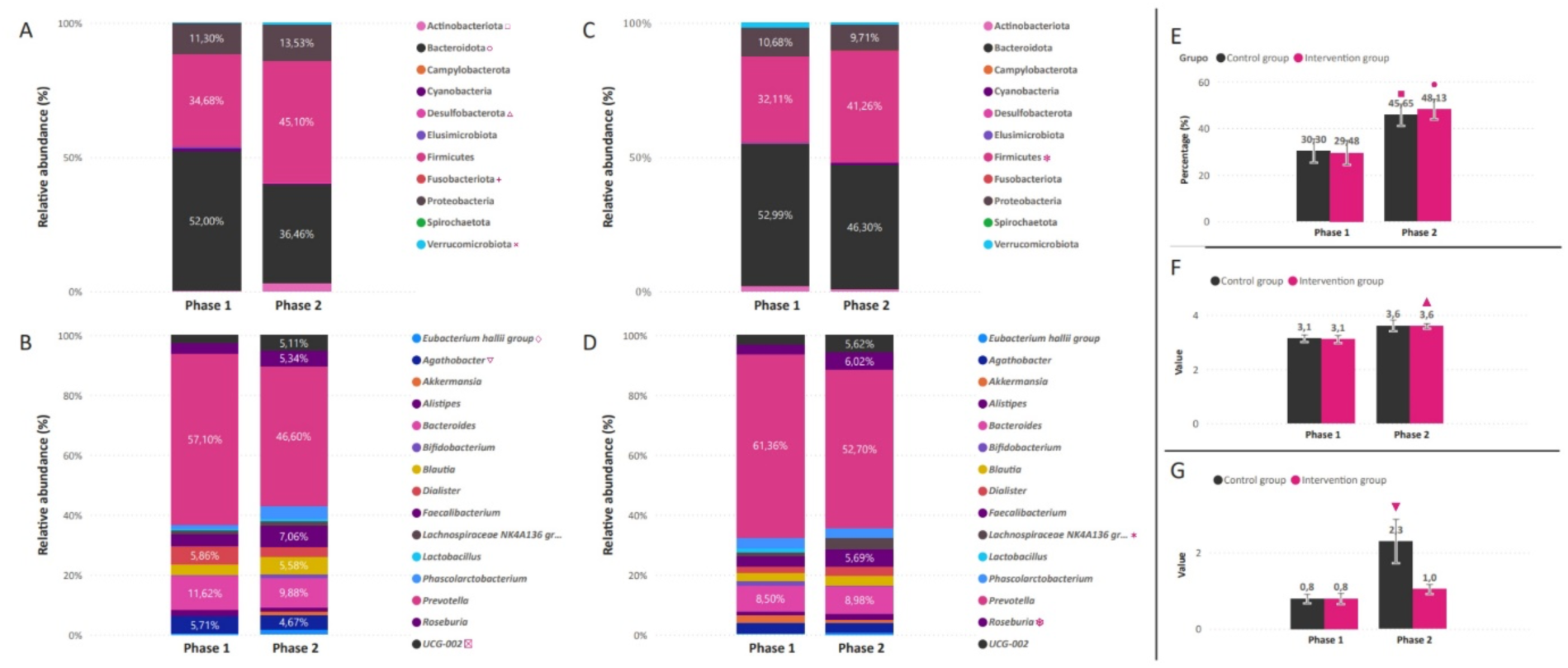
Changes in gut microbiome biomarkers during the study. **(A)** Relative abundance of bacterial phyla in the CG across both study phases. **(B)** Relative abundance of the most representative bacterial genera in the CG. **(C)** Relative abundance of bacterial phyla in the IG. **(D)** Relative abundance of the most representative bacterial genera in the IG. **(E)** Percentage of the Biomatest gut health index for both phases in the control and IG **(F)** Shannon index values for both groups across study phases.**(G)** F/B ratio values in both phases for the control and IG. Pink symbols indicate food groups with statistically significant changes, determined by a nonparametric Kruskal-Wallis analysis at a significance level of p < 0.05. Significant changes in bacterial groups between phases in the CG include increases in *Eubacterium hallii* (♢, p-value = 0.04), *UCG-002* (☒, p-value = 0.03), *Actinobacteriota* (□, p-value = 0.00), *Desulfobacterota* (△, p-value = 0.01), *Fusobacteriota* (+, p-value = 0.04), and *Verrucomicrobiota* (×, p = 0.04), as well as a decrease in *Agathobacter* (▽, p = 0.02) and *Bacteroidota* (○, p = 0.01) (ANOVA) (Figure 3 A, B).In the IG, a statistically significant increase was observed between phases in *Lachnospiraceae_NK4A136_group* (✶, p-value = 0.01), *Roseburia* (⎈, p-value = 0.03), and Firmicutes (✽, ANOVA p-value = 0.05). The intestinal health index (▪, p-value = 0.02) and F/B ratio (●, p-value = 0.02) also showed significant differences. Furthermore, for the IG, significant changes were noted in the Shannon index (▴, ANOVA p-value = 0.01) and the intestinal health index (▾, p-value = 0.01) (Figure 3 E, F, G).

**Figure 3.**
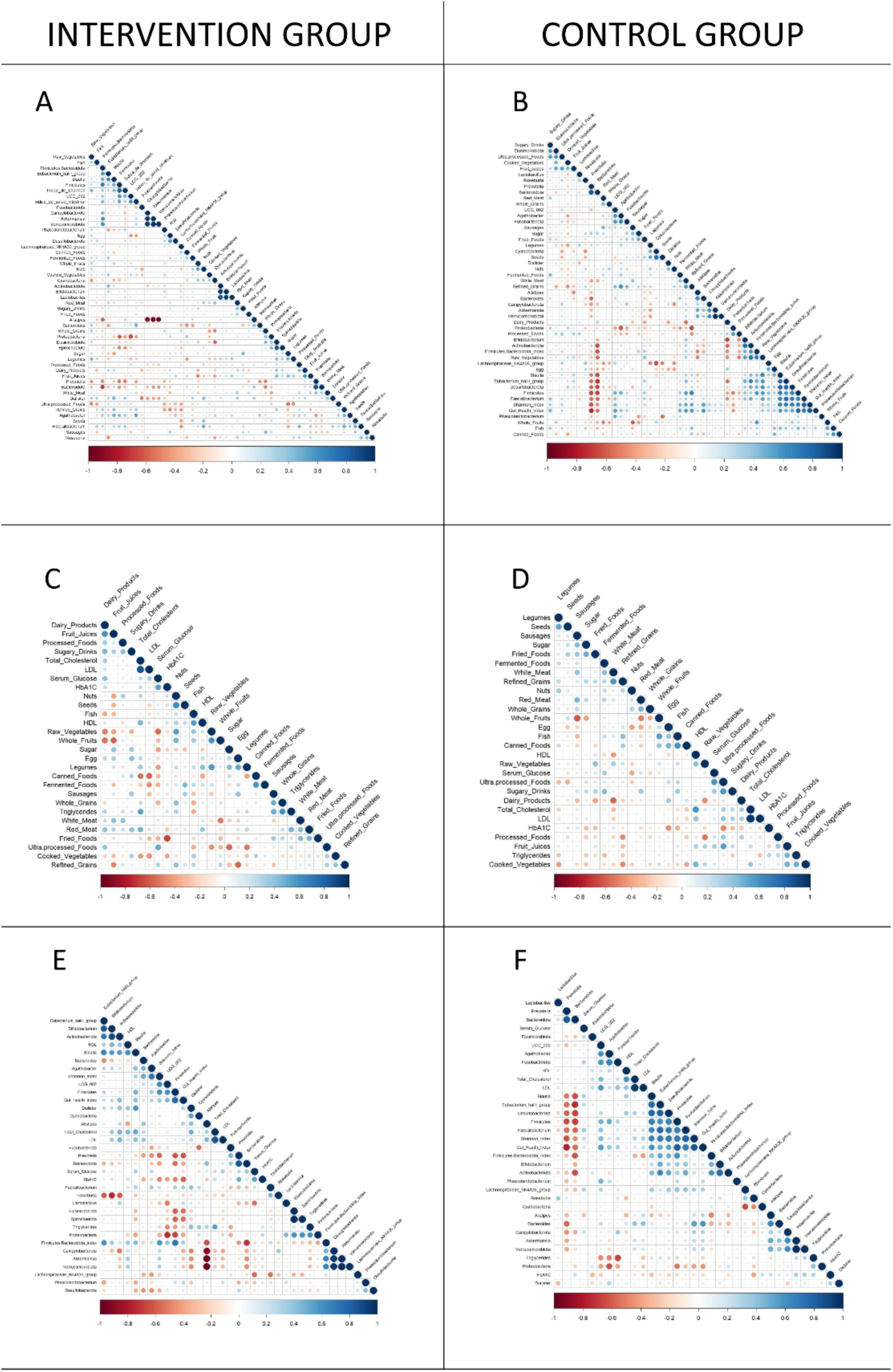
Correlation of blood, gut microbiome and food intake biomarkers. Figures (A), (C), and (E) show correlations for the between (A) changes in microbiome indicators and food group changes, (C) changes in blood chemistry biomarkers and food group changes, and (E) changes in blood chemistry biomarkers and microbiome indicators. Panels (B), (D), and (F) display the same correlations for the CG. Statistically significant correlations are highlighted…

### There are significant correlations between changes in microbial groups and gut microbiota indicators, blood chemistry biomarkers, and changes in food consumption frequencies

Negative correlations (which means there is an inverse proportional increase between both elements, when one increases, the other decreases) were found between serum glucose and raw vegetables (-0.56) (Fig. 3B), serum glucose and dairy products (0.42) (Fig. 3E). Positive correlations (which means there is a directly proportional increase between both elements, when one increases, the other also increases) were found between serum glucose and ultra-processed foods (0.5) (Fig. 3E)., with total cholesterol, and red meat (0.46) (Fig. 3B). For LDL cholesterol, a negative correlation was found with fermented foods (-0.47), and for triglycerides, a positive correlation was found with processed meats (0.5) and fruit juices (0.44) (Fig. ure3B). Additionally, refined grains consumption had a negative correlation with stress (-0.43), anxiety (-0.50), and depression (-0.47) (results not shown in Figures; shown in Supplementary Material 6 Fig. S1.)

For correlations regarding blood chemistry and bacterial groups, negative ones were found between serum glucose and *Lactobacillus* (-0.54) and *Lachnospiraceae NK4A36 group* (-0.56) (Fig. 3C). A positive correlation was found between total cholesterol and *Dialister* (0.5), LDL cholesterol and *Proteobacteria* (0.59), and a negative correlation between triglycerides and *Agathobacter* (-0.48) and *UCG-002* (-0.48).

Correlations between bacterial groups and foods were found for between *Faecalibacterium* and nuts (0.49), seeds (0.49), and cereals (0.49). A positive correlation was found between *Eubacterium hallii* group and raw vegetables (0.6). *Blautia* showed a positive correlation with raw vegetables (0.62) and eggs (0.4). *Desulfobacteriota* also had a positive correlation with eggs (0.56), as did *Lachnospiraceae* NK41 (0.5). A negative correlation was found between *UCG-002* and eggs (-0.45), and for *Bifidobacterium*, a positive correlation was found with raw vegetables (0.5) and a negative correlation with dairy products (-0.6).

There were also correlations found between bacterial groups themselves: a positive correlation between *Bifidobacterium* and *Lactobacillus* (0.94) and a negative correlation between *Alistipes* and *Verrucomicrobiota* (-0.94). Correlations found between bacterial groups and intestinal indices: positive correlations were found between *Faecalibacterium* and Shannon Index (0.81), and between *Eubacterium hallii* group and Shannon Index (0.69), Simpson Index (0.49), and F/B ratio (0.51). Negative correlations were found between *Prevotella* and Shannon Index (-0.64), and between *Alistipes* and F/B Index (-0.49).

## Discussion

Our study examines the impact of personalized interventions in diet and lifestyle on the gut microbiome of people with obesity diagnosis, based on their initial microbiome composition. The general hypothesis suggests that generating and following personalized dietary recommendations based on the microbiome, in participants with this condition (as well as other conditions), lead to positive changes in the gut microbiome, promoting taxa associated with improved metabolic health.^44, 45, 46^ This aligns with the widely documented fact that the gut microbiome is dynamic and responds to changes in diet and lifestyle,^44, 47, 48^ potentially impacting in a direct way through various metabolic pathways involved in immunity, energy, lipid and glucose metabolism, the treatment of people with overweight and obesity.^49^ The results demonstrate, in significant values within the sample of the population of study (cohort) a complex interaction between dietary choices, gut microbial composition, and metabolic health, highlighting the importance of addressing not only weight management but also the inflammation associated with obesity through a dysbiotic microbiome.^50, 51, 52^

The study shows an unexpected abundance of Bacteroidota, specifically of the genera *Bacteroides* and *Prevotella* in the cohort of participants with obesity diagnosis, which were mostly characterized by unhealthy eating habits, especially in the phase one of this study, suggesting that dietary components can directly influence the microbiota.^8, 53, 54^ and modulate the relative abundance of groups such as Bacteroidota or Enterobacteriaceae, which have been previously described as diminished^55, 56^ or increased^49, 57^ respectively, in other studies in people and mice with this condition. The high prevalence of Proteobacteria (from 9.5-13.3% relative abundance, Fig. 2A y 2C) in participants with obesity diagnosis in this cohort, as well as its increase in the CG and its decrease in the treated group (Fig.2A and 2C) underscores its potential link to chronic inflammation,^58^ consistent with recent studies which find it increased in obesity populations.^59, 60^ Additionally, after the intervention, we observed a significant increase in *Eubacterium hallii* and *UCG-002* in the IG, both known for their protective effects on gut health, as well as the *Roseburia* genus and microbial diversity.^38, 39, 61, 62^ The CG, which did not receive microbiome-based recommendations, showed a decrease in the beneficial *Agathobacter* genus, which has been related to anxiety and sleep problems.^63, 64^ The fact that there was no significant weight loss in the cohort between the two phases, but also no weight gain, and that almost a kilo and a half was lost through microbiome-focused interventions, is an important finding of this study.^65, 66, 67^

The distinctive role played by tests based in metagenomics that diagnose and not only assess but analyze the microbiome, not only intestinal but also vaginal and oral, has become evident in their ability to offer a comprehensive evaluation of gut health, particularly in this study.^68^ As we seek to understand a patient’s diet and lifestyle, microbiome modulation, and blood chemistry biomarkers, these tests emerge as a precise and relevant tool, extending beyond research into clinical practice, valuable for understanding and improving gut health in the context of obesity. Although in their early stages and still with limited capacity to establish direct correlations with the patient’s clinical outcome, they enable a comprehensive evaluation of the impact of personalized interventions on the microbiome, inflammation, dietary preferences, and the adoption of new habits. The low values found in the initial measurement of the Gut Health Index as measured by the test used in this study, suggest a dysbiosis state associated with obesity.^65, 69^

In this study, we rigorously aimed to generate dietary interventions that could modulate the microbiome, with this being the focus rather than caloric restriction or weight loss.^70, 71^ As a background, the Mediterranean diet (MD) has been recognized as an effective regulator of the gut microbiome due to its emphasis on plant-based foods and moderate consumption of animal products. Adherence to the MD is inversely associated with chronic disorders such as obesity and type 2 diabetes. Long-term adherence to the MD has been shown to increase *Bifidobacterium* and *Bacteroides* while decreasing Firmicutes.^72, 73^ This reinforces the importance of plant-based diets for promoting a healthy gut microbiota. Therefore, our intervention represents a novel, affordable food grouping for populations in countries with a high proportion of low-income people, such as Colombia,^74^ that can be extended to other Latin American countries. It includes culturally appropriate foods like plantains, bananas, and corn, and incorporates evidence presented here, that improves the gut microbiome. This finding is also relevant for other populations at risk of malnutrition and with serious implications, such as children and pregnant women in vulnerable and low-income populations (Article on microbiome and malnutrition in Africa).^75^ This is relevant in the light that the gut microbiome shows remarkable resistance to most temporary external influences, although the overall microbial community exhibits high inter-individual variability.^76^ However, one of the most impactful factors on it is food and changes in food frequencies^45, 77^ Intestinal microorganisms are continuously and extensively renewed, with the ability to double in number within an hour, and it is believed that rapid changes in microbial composition at the species and family levels occur within 24-48 hours after a dietary intervention.^78^ Similarly, mouse models have shown that manipulating macronutrient intake consistently changes gut microbiome composition within a day.^38, 39, 61, 62^ Despite these observed changes, the fundamental question remains whether modifications to the gut microbiome will persist depending on the duration of the dietary intervention.

The second phase of our study provided valuable insights into the temporal aspects of microbiome modulation. We observed changes in the weekly intake frequencies of food groups (Fig. 2), indicating a significant habit change (increase) in the frequency of cereal and seed consumption and a decrease in dairy, table sugar, and ultra-processed foods in the IG between Phases 2 and 1 of the study. This aligns with the intervention, which, as detailed in the methodology, was based on the findings of the microbiome composition of the IG during Phase 1, directly and repeatedly recommending an increase in fiber through legumes, whole grains, tubers, fruits, vegetables, nuts, and seeds. A decrease in ultra-processed and high-sugar foods was also recommended. The CG results were contrary, with no precise or repetitive microbiome-based intervention, where only a significant decrease in processed food consumption was observed. This may be due to the bias participants experience just by being part of a nutrition-related study, where processed foods are among the most restricted food groups. Regarding microbiome composition between study phases 1 and 2 (Fig. 3), increases were observed in *Eubacterium hallii* (♢ p-value = 0.04211) and *UCG-002* ( p-value = 0.03219), two bacteria known for their protective effects on the microbiome and indicators of good gut health,^79, 80, 81, 82, 83^ which in this study may have increased as processed foods decreased. A decrease in *Agathobacter* was observed in the CG (Fig. 3B), which may signify an increase in anxiety and a reduction in sleep quality, as this group has been reported as involved in these processes.^84, 85^ In the IG, there was a significant increase in the *Roseburia* genus (Fig. 3B) and microbial diversity (Fig. 3F), both indicators of increased fiber and prebiotic foods.

Finally, correlations validated by the literature and novel correlations were identified, which can guide future dietary interventions. Among the validated ones, we found a positive correlation between total cholesterol and red meat (0.46), as well as with the genus *Dialister* (0.5).^86, 87, 88, 89^ Triglycerides also correlated positively with processed meats (0.5) and serum glucose with ultra-processed foods (0.5).^90, 91, 92, 93, 94^ Likewise, the consumption of raw vegetables was negatively correlated with serum glucose (-0.56) and positively associated with *Lactobacillus* and *Lachnospiraceae*, both known for their role in improving gut health.^95, 96, 97, 98, 99, 100^. It is widely reported, especially for the first taxonomic group, that it positively impacts metabolism, meaning a decrease in serum glucose levels. Some species involved in these glucose-lowering roles are: *Lactobacillus rhamnosus* BSL; La*ctobacillus rhamnosus* R23, and strains of *L. reuteri* ADR-1 and ADR-3.^98, 99, 100^ Serum glucose also had a positive correlation with dairy (0.42) (Fig. 3). In other studies,^95^ there was a significant and substantial relationship between dairy consumption and insulin resistance. Contrary is the serum glucose consumption with raw vegetables, which show a negative correlation (-0.56) (Fig. 3).^101, 102^ Previous research reports that salad and raw vegetable consumption are associated with a reduced risk of abnormal glucose tolerance.^96, 97^

In terms of novel, not previously reported correlations, we found that, LDL negatively correlateds with fermented foods (-0.47) and positively with *Proteobacteria* (0.59), a phylum linked to metabolic diseases and dysbiosis.^103, 104^ Increased relative abundance of *Proteobacteria* has been reported in patients with chronic cardiac syndrome.^105, 106^ A high relative abundance of this phylum (*Proteobacteria*¿ is associated with gut dysbiosis and other metabolic diseases, including cancer.^104^ Other correlations included the positive relationship of triglycerides with fruit juice consumption (0.44) and the negative correlation with *Agathobacter* and UCG-002 (-0.48), a group of bacteria found to protect the gut microbiome.^107, 108, 109, 110^ Correlations were also found with psychological factors, consistent with the described relationship between gut health and mental health,^111, 112^ specifically finding that higher cereal consumption correlates negatively with stress (-0.43), anxiety (-0.50), and depression (-0.47) (Supplementary Material 6, Figure S1.), denoting the positive impact of a cereal-rich diet on mental health. Particularly, the genus *Faecalibacterium* showed a positive correlation with nuts, seeds, and cereals (0.49), fiber-rich foods, suggesting an improvement in microbial diversity.^113^ Conversely, *Prevotella* negatively correlated with gut diversity, which could imply adverse effects on mental health.^114, 115^

Specific correlations of microorganisms with undesirable effects on the gut microbiome were also evidenced. *Prevotella* negatively correlates with the Shannon Index or gut diversity (-0.64).^81^ The phylum Desulfobacteriota correlates positively with egg consumption (0.56) in this study. We hypothesize, that due to this food being high in proteins and sulfur-containing amino acids such as Cysteine and methionen, these bacteria (Desulfobacteriota) can use them as substrate and convert them to hydrogen sulfide. Although, there has been recent evidence on the beneficial effects of egg consumption in the gut microbiota.^116, 117, 118^ On the contrary, the genus UCG-002 negatively correlated with eggs (-0.45), indicating that the intake of this food, generated compounds that negatively affect this bacterium. The genus *Alistipes*, widely linked to states of stress, anxiety, and mood disorders,^116^ negatively correlates with the F/B ratio (-0.49), which is described in the literature.^114^ This cited study, conducted with German participants with obesity diagnosis, found that *Alistipes* decreases with obesity. Another study^115^ found that *Alistipes* was more abundant at baseline in a weight loss program, with patients who had a higher relative abundance of this microorganism being more effective and sustainable in weight loss.

The genera *Blautia* positively correlated with egg consumption (0.4 and 0.56 correlation index, respectively) (Fig. 3). In previous studies on young and middle-aged rats, the percentages of relative abundance of *Blautia* increased after consuming eggs for 14 days.^119,120^ *Blautia* also positively correlates with raw vegetables (0.62) (Fig. 3). Its effect is widely supported by the prebiotic compounds contained in raw vegetables, especially polyphenols. Studies have shown that polyphenols exert prebiotic effects by selectively stimulating the growth of beneficial microorganisms, including species belonging to the genera *Lactobacillus, Bifidobacterium, Akkermansia, Roseburia, Ruminococcus,Blautia, Dorea, and Faecalibacterium*. Due to their potential benefits, a diet rich in polyphenol-containing foods such as fruits, vegetables, green tea, coffee, red wine, and dark chocolate is recommended.^121,122, 123^ A negative correlation was found between the genus *Alistipes* and the phylum Verrucomicrobiota (-0.94) (Fig. 3), which could be explained by the fact that this genus has been linked to dysbiosis and mental health conditions such as anxiety and depression. Meanwhile, the phylum Verrucomicrobiota, with *A. muciniphila* as a representative, is related to the synthesis of neuroactive metabolites (short-chain fatty acids, GABA, among others) that decrease the occurrence of mental health conditions.^124, 125^ The *Eubacterium hallii* group positively correlated with the Shannon Index (0.69), indicator of gut diversity, and the F/B Index (0.51), an indicator of metabolic balance of the microbiota.^126, 127^ A healthy gut microbiome generally includes a good proportion of butyrate-producing microorganisms, such as *Eubacterium hallii*, contributing to a stable Firmicutes/Bacteroidota index.^128, 129^

In conclusion, this study constitutes a significant contribution to the growing body of knowledge around the complex relationship between diet, the microbiome, and metabolic health. Some of the observed variations in the variables of changes in food group consumption and gut microbiome composition were significant, especially in the IG, which received precision recommendations based on the microbiome, with systematic follow-up. This denotes effectiveness in obesity treatments, especially in habit retraining, by using diagnostic tools such as microbiome tests based on detecting microbial composition through metataxonomy or 16S rRNA gene sequencing. Changes were also observed in the measured variables for the controls, which are explained considering a behavior change biased by belonging to a study, which might induce the improvement of certain habits.^101, 102^Changes in blood chemistry biomarkers were not significant in either of the two study groups, demonstrating that a longer adherence period to the indicated dietary changes is required to improve the microbiome, so that these effects in gut bacteria are reflected in blood biomarkers. Still, the findings allow evidence of statistically significant correlations between the abundances of certain microbial groups with blood biomarkers and changes in the frequency of consumption of certain food groups, opening a window of possibility for the modulation of variables such as blood glucose, total cholesterol, LDL cholesterol, and total triglycerides through modulation of the gut microbiome, which in turn is modulated by changes in food groups.

## Data Availability

All data produced in the present study are available upon reasonable request to the authors

## Author Contributions

All authors contributed to the study conceptualization and manuscript writing. SBJ, IAR, VCM, LGM, JDST, and MCAE were responsible for volunteer contact and follow-up, while SBJ, IAR, VCM, and LGM conducted the experiments and assay development. The methodology and statistical analysis were carried out by CAZ, LSZ, AM, LGM, SLO, MCAE, SBJ, IAR, and VCM, with computational code developed by SLO and SBJ. Visualization of results was handled by VCM and SBJ. AM, JDST, LGM, LSZ, and MCAE provided leadership and supervision, and project administration and resources were managed by JDST, LSZ, AM, and LGM. All authors reviewed and approved the final manuscript.

## Acknowledgments

We thank Comfama for their trust and funding, which enabled this study and supported health initiatives in Colombia. Our appreciation also goes to the Comfama contact team for their dedication in reaching out to participants and ensuring smooth coordination.

We extend our gratitude to the Astrolab Bio team for their expertise in analyses, personalized recommendations, and participant follow-ups (Natalia Betancur and Verónica Pérez). Additionally, we acknowledge the physicians for their key role in ensuring the relevance of recommendations and effectively presenting the results.

Finally, we thank all participants and contributors who supported this project, helping advance precision nutrition in our country.

