## Supplemental Material 2 for "Low-Cost Precision nutrition recommendations, generated by metataxonomy-based microbiome tests, improve food group choices and gut health indicators in a population with obesity diagnosis in Colombia"

**Inclusion and exclusion criteria**

- Persons belonging to the SURA-COMFAMA database.
- People between 25 and 55 years of age.
- Persons with a Body Mass Index (BMI) greater than or equal to 30.
- Residents of Medellín or municipalities in the Aburrá Valley, Antioquia.
- For women: Not pregnant or planning to become pregnant within the next 9 months.
- For women: Not be breastfeeding.
- Not be in the process of bariatric surgery or obesity treatment surgery or have already undergone this type of intervention.
- Not be on a restrictive diet or on a specific nutritional plan.
- Not be diagnosed with a mental illness that requires psychiatric accompaniment.
- Do not plan to travel in the next 9 months. If you do, be willing to inform the team.

**Not having consumed any of the following medications during the last three months prior to participation in the study:**

- Valproic Acid (Anticonvulsant, Chronic Migraines, TAB)
- Antihistamines/Antiallergics
- Amitriptyline (Tricyclic Antidepressant, Anxiety disorders and insomnia)
- Atorvastatin (Hypercholesterolemia)
- Azithromycin (Antibiotic for respiratory infections)
- Ceftazidime/Avibactam (Broad-spectrum antibiotic)
- Diclofenac (NSAID)
- Doxycycline (Antibiotic in soft tissue and skin infections)
- Oral steroids (Hydrocortisone, dexamethasone, prednisone) for Inflammatory and rheumatological diseases
- Fluoxetine, Escitalopram, Venlafaxine or Duloxetine (Antidepressant, anxiolytic, hypnotic and sedative)
- Laxatives
- Losartan (Antihypertensive ARA 2)
- Metformin (Hypoglycemic in the treatment of DM)
- Metoprolol (Beta-blocker in cardiac arrhythmias, Antihypertensive)
- Metronidazole (Antiparasitic)
- Omeprazole (PPI)
- Oxycodone (Opioid for the treatment of chronic pain)
- Paracetamol/Acetaminophen (NSAID analgesic action)
- Propofol (Sedative and Hypnotic)
- Ritonavir (Antiretroviral for HIV management)
- Topiramate, Carbamazepine, Lacosamide, Lamotrigine (Anticonvulsant in refractory and non-refractory epilepsies, absence seizures).
- Quetiapine (mood stabilizer, TAB, treatment of mania)
- Warfarin (Mechanical Valvular Prosthesis)
