## Supplemental Material 5 for "Low-Cost Precision nutrition recommendations, generated by metataxonomy-based microbiome tests, improve food group choices and gut health indicators in a population with obesity diagnosis in Colombia"

**BIOMATEST Follow-up Survey (Intervention Group)**

1. E-mail address

2. Full name

3. In the last 15 days, have you taken antibiotics?

4. In the last 15 days have you travelled for more than a week?

5. In the last 15 days, have you had any surgical interventions?

6. In the last 15 days, have you started taking any medication?

7. In the last 15 days, how many hours of sleep per day on average have you had?

8. In the last 15 days, has your sleep been restful (restful = having energy, not feeling fatigue and carrying out daily activities in a normal way)?

9. In the last 15 days, how often have you been physically active?

10. Which of the following physical activities have you done?

Cardiovascular (walking, jogging, spinning, cycling, elliptical, swimming, dancing, aerobics)

Strength (weight training, weights)

Cardiovascular + strength

None

11. How long do you exercise for each session?

12. In the last 15 days, how often have you eaten legumes (e.g. beans, lentils, chickpeas, blanquillos, kidney beans, peas)?

13. How often have you eaten whole fruits in the last 15 days (not counting juices or smoothies)?

14. How often have you eaten raw or cooked vegetables in the last 15 days?

15. In the last 15 days how often have you eaten whole grains (e.g. brown rice, whole grain pasta, rolled oats, quinoa, whole grain bread, natural corn on the cob, rye)?

16. In the last 15 days how often have you consumed fermented foods (e.g. sauerkraut, milk kefir, water kefir, yoghurt, kombucha)?

17. In the last 15 days how often have you consumed healthy fats (e.g. olive oil, ghee, coconut oil, avocado oil)?

18. In the last 15 days how often have you eaten processed foods (examples: fast foods, crackers, toast, cookies, white bread, white or yellow pasta, white rice)?

19. In the last 15 days, how often have you eaten ultra-processed foods (examples: packaged foods, energy drinks, boxed cereals, sachet soups)?

20. How often have you consumed alcoholic beverages in the last 15 days?

21. In the last 15 days, how often have you consumed added sugar in your meals?

22. In the last 15 days, how often have you eaten fried foods?

23. How often per week did you eat the following colors of the food rainbow?

[Red]

[Orange]

[Yellow]

[Green]

[Blacks and Purples]

[Brown]

[White]

24. Do you feel full after each main meal of the day?

25. In the last 15 days, how would you rate your compliance with the diet and lifestyle recommendations on a range of 0 to 10?

Please read the following statements and indicate from 0 to 3 to what degree this statement has happened to you during the past week. The rating scale is as follows: 0: It has not happened to me; 1: It has happened to me a little, or some of the time; 2: It has happened to me a lot, or a good part of the time; 3: It has happened to me a lot, or most of the time.

I had a hard time releasing the tension

I realized that my mouth was dry

I couldn't feel any positive feelings

I found it difficult to breathe

I found it difficult to take the initiative to do things

I overreacted in certain situations

I felt my hands shaking

I felt that I was expending a great deal of energy.

I was worried about situations in which I might panic or where I might make a fool of myself.

I have felt that there was nothing to look forward to.

I felt restless

I found it difficult to relax

I felt sad and depressed

I did not tolerate anything that did not allow me to continue with what I was doing.

I felt I was at the point of panic

I couldn't get excited about anything

I felt I was worth very little as a person

I have tended to feel angry easily

I felt my heart pounding even though I had not made any physical exertion

I was afraid for no reason

I felt that life had no meaning

**BIOMATEST Monitoring Survey (Control Group)**

1. E-mail address

2. Full name

3. In the last 15 days, have you taken antibiotics?

4. In the last 15 days have you travelled for more than one week?

5. In the last 15 days have you had any surgical interventions?

6. In the last 15 days, have you started taking any medication?

7. In the last 15 days, how many hours of sleep per day on average have you had?

8. In the last 15 days, has your sleep been restful (restful = having energy, not feeling fatigue and carrying out daily activities in a normal way)?

9. In the last 15 days, how often have you been physically active?

10. Which of the following physical activities have you done?

Cardiovascular (walking, jogging, spinning, cycling, elliptical, swimming, dancing, aerobics)

Strength (weight training, weights)

Cardiovascular + strength

None

11. How long do you exercise for each session?

12. In the last 15 days, how often have you eaten legumes (e.g. beans, lentils, chickpeas, blanquillos, kidney beans, peas)?

13. How often have you eaten whole fruits in the last 15 days (not counting juices or smoothies)?

14. How often have you eaten raw or cooked vegetables in the last 15 days?

15. In the last 15 days, how often have you eaten whole grains (e.g. brown rice, whole wheat pasta, rolled oats, quinoa, whole grain bread, natural corn on the cob, rye)?

16. In the last 15 days how often have you consumed fermented foods (e.g. sauerkraut, milk kefir, water kefir, yoghurt, kombucha)?

17. In the last 15 days how often have you consumed healthy fats (e.g. olive oil, ghee, coconut oil, avocado oil)?

18. In the last 15 days, how often have you eaten processed foods (examples: fast foods, crackers, toast, cookies, white bread, white or yellow pasta, white rice)?

19. In the last 15 days, how often have you eaten ultra-processed foods (examples: packaged foods, energy drinks, boxed cereals, sachet soups)?

20. How often have you consumed alcoholic beverages in the last 15 days?

21. In the last 15 days, how often have you consumed added sugar in your meals?

22. In the last 15 days, how often have you eaten fried food?

23. Do you feel full after each main meal of the day?

Please read the following statements and indicate from 0 to 3 to what degree this statement has happened to you during the past week. The rating scale is as follows: 0: It has not happened to me; 1: It has happened to me a little, or for some of the time; 2: It has happened to me a lot, or for a good part of the time; 3: It has happened to me a lot, or most of the time.

I had a hard time releasing tension

I noticed that my mouth was dry

I could not feel any positive feelings

I found it difficult to breathe

I found it difficult to take the initiative to do things.

I overreacted in certain situations

I felt my hands shaking

I felt that I was expending a great deal of energy.

I was worried about situations in which I might panic or where I might make a fool of myself

I have felt that there was nothing to look forward to.

I felt restless

I found it difficult to relax

I felt sad and depressed

I did not tolerate anything that did not allow me to continue with what I was doing.

I felt I was at the point of panic

I couldn't get excited about anything

I felt I was worth very little as a person

I have tended to feel angry easily

I felt my heart pounding even though I had not made any physical exertion

I was afraid for no reason

I felt that life had no meaning
