## Supplementary figures and images for "Low-Cost Precision nutrition recommendations, generated by metataxonomy-based microbiome tests, improve food group choices and gut health indicators in a population with obesity diagnosis in Colombia"

### Supplemental Material 6

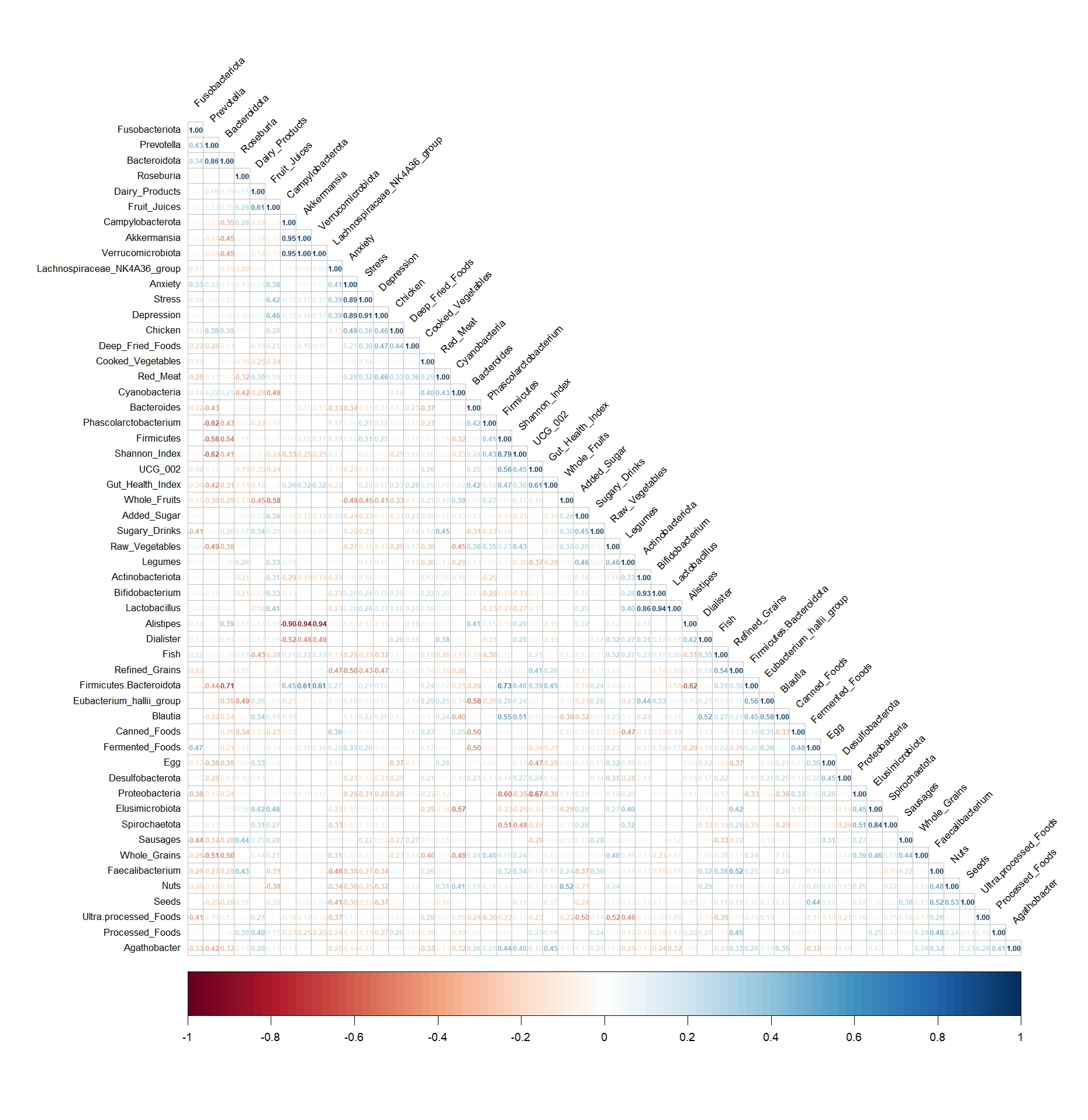

**Figure 1. Correlation of blood, gut microbiome, food intake and mental health biomarkers.**
